# Knowledge, attitude and perception regarding HIV/AIDS among Khartoum University medical campus students – a cross-sectional study 2018

**DOI:** 10.1101/2024.08.23.24311187

**Authors:** Ahmed Alasaad Kamel Mohamed Elamir, Rana Ibrahim Abdalla Dafalla, Raghda Ali, Mohamed O. Elhussain, Ahmed Abdelaziz Abugrain Abdelsalam, Abraa Awad Sharif, Sayed Halali

## Abstract

**Background:** Clinical training is a part of curriculum for medical campus students, where there is a chance of exposure to HIV infection. Hence, this study was conducted to assess their knowledge, attitude and perception towards HIV/AIDS.

**Method:** This is a descriptive, non-interventional, cross-sectional study. Data collection took place December 2017 and January 2018. The study population included medical students at the University of Khartoum during the study period. Data were collected through a structured adapted questionnaire covering socio demographic details, General knowledge about HIV/AIDS, medical campus students perception towards the possible causes of HIV infection, medical campus students attitude towards HIV/AIDS patients used to collect data from medical students.

**Results:** This study investigated the knowledge, attitude and perception regarding HIV/AIDS among 288 medical students at University of Khartoum. It was found that the means and standard deviation of the total knowledge, attitude and perception score towards HIV/AIDS in general were 15.45±2.93. which was categorized fair (reference ranging from 13 to 18). the total knowledge, attitude and perception was significantly associated with the academic year (p=0.000), but no association was found between the attitude score and the academic year (p = 0.620).

**Conclusion:** The overall rate of knowledge and attitude about HIV/AIDS among medical campus students of Khartoum University was acceptable. However, appreciated number of the students have had a negative attitude towards HIV/AIDS patient.

## Background

The Human Immunodeficiency Virus (HIV) targets the immune system and weakens people’s defence systems against infections and some types of cancer. As the virus destroys and impairs the function of immune cells, infected individuals gradually become immunodeficient. Immune function is typically measured by CD4 cell count. Immunodeficiency results in increased susceptibility to a wide range of infections, cancers and other diseases that people with healthy immune systems can fight off.

The most advanced stage of HIV infection is Acquired Immunodeficiency Syndrome (AIDS), which can take from 2 to 15 years to develop depending on the individual. AIDS is defined by the development of certain cancers, infections, or other severe clinical manifestations.**(1)**

The Acquired Immunodeficiency Syndrome (AIDS) epidemic is in its third decade and has become a pandemic disease that threatens the world population. It affects all body systems as well as the mental health and social relationships of carriers and asymptomatic patients. The human immunodeficiency virus (AIDS) and the acquired immunodeficiency syndrome (AIDS) have profoundly affected every aspect of the public health sector.**(2)** Knowledge about the spread of HIV and safe sexual practices has a critical impact on the prevention of the acquired immunodeficiency syndrome (AIDS). Sexually Transmitted Infections (STI’s), including HIV (Human Immunodeficiency Virus) mainly affects sexually active young people. Young adults aged 15–29 years, account for 32% of AIDS (Acquired Immunodeficiency Syndrome). Causes of the increased rates of STIs/HIV in young people are complex, however, the main reasons include biological factors, risky sexual behaviour patterns (early initiation of sex, premarital sex, bisexual orientation and multiple sexual partners), transmission dynamics and treatment-seeking behaviour. There is growing evidence of increased premarital sexual activities among young people. While generalisation is difficult, studies indicate that between 20% and 30% of young men and up to 10% of young women have premarital sexual experiences. Women, have a higher incidence of HIV infection than men because of their greater biological susceptibility **(3)**.

The symptoms of HIV vary depending on the stage of infection. Though people living with HIV tend to be most infectious in the first few months, many are unaware of their status until later stages. The first few weeks after initial infection, individuals may experience no symptoms or an influenza-like illness including fever, headache, rash, or sore throat. As the infection progressively weakens the immune system, an individual can develop other signs and symptoms, Such as swollen lymph nodes, weight loss, fever, diarrhoea and cough. Without treatment, they could also develop severe illnesses such as tuberculosis, cryptococcal meningitis, severe bacterial infections and cancers such as lymphomas and Kaposi’s sarcoma, among others.**(1,4)**.

HIV can be transmitted via the exchange of a variety of body fluids from infected individuals, such as blood, breast milk, semen and vaginal secretions. Individuals cannot become infected through ordinary day-to-day contact such as kissing, hugging, shaking hands, or sharing personal objects, food or water.**(1)**.

Risk factors for contracting HIV includes: having unprotected anal or vaginal sex, having another sexually transmitted infection (;such as syphilis, herpes, chlamydia, gonorrhoea, and bacterial vaginosis), sharing contaminated needles or syringes and other injecting equipment and drug solutions when injecting drugs, receiving unsafe injections, blood transfusions, tissue transplantation, medical procedures that involve unsterile cutting or piercing; and experiencing accidental needle stick injuries, including among health workers.**(1,6,8)**.

## Methods

### Study setting

This study was conducted in the medical campus, university of Khartoum, at El Qasr Ave.

### Study design

A descriptive cross-sectional study was conducted in the medical campus, University of Khartoum between December 2017 and January 2018. An author designed closed-ended questionnaire (25items) consisting of socio demographic details, General knowledge about HIV/AIDS, medical campus students perception towards the possible causes of HIV infection, medical campus students attitude towards HIV/AIDS patients used to collect data from medical students. The number of participants from each batch was determined according to batch percentage from all campus population. inside the batch convenient random sampling technique used to choose the students under the study.

### Inclusion and exclusion criteria

The sample was obtained from second and fifth year medical students, second and fifth year Dentistry students and second and fifth year pharmacy students, whom agreed to participate in this study.

### Sample size

Sample size has been calculated according to following formula:

n=z2 pq/d2,when ;

n= sample size.

z= the normal standard deviation (z=1.96).

p= the prevalence; (The prevalence in Sudan is 0.25)**(12)**

q= 1-p (the frequency of non occurrence of an event) = 1- 0.25= 0.75

d= level of precision (= 0.05)

Then the sample size has found to be 288. Refer to Table 1 to view the sample selection and response rates.

**Table (1):** shows the mean knowledge, attitude, perception and the final knowledge, attitude and perception score.

| Statistics |  |  |  |  |
| --- | --- | --- | --- | --- |
|  | KSCORE | PSCORE | ASCORE | Final_KAP |
| Valid | 263 | 271 | 273 | 262 |
| Missing | 14 | 6 | 4 | 15 |
| Mean | 7.9125 | 5.4760 | 2.0806 | 15.4542 |
| Median | 8.0000 | 6.0000 | 2.0000 | 16.0000 |
| Std. Deviation | 1.69304 | 1.20122 | 1.22509 | 2.93542 |
| Range | 7.00 | 5.00 | 4.00 | 14.00 |
| Minimum | 3.00 | 2.00 | .00 | 7.00 |
| Maximum | 10.00 | 7.00 | 4.00 | 21.00 |

**Table (2):** Illustrates the association between the knowledge score and academic year.

**KAP\_SCORE2 \* Academic year Crosstabulation**
| Count |  |  |  |  |
| --- | --- | --- | --- | --- |
|  |  | Academic year |  | Total |
|  |  | Second year | Fifth year |  |
| KAP_SCORE2 | POOR | 28 | 9 | 37 |
|  | FAIR | 79 | 101 | 180 |
|  | GOOD | 10 | 31 | 41 |
| Total |  | 117 | 141 | 258 |

**Table (3):**
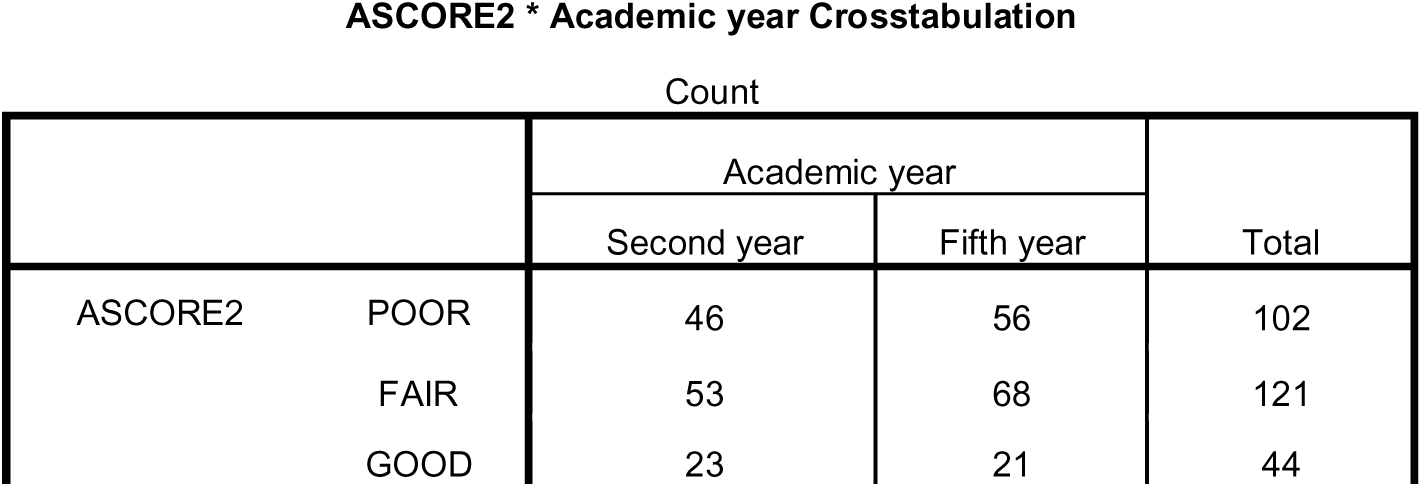
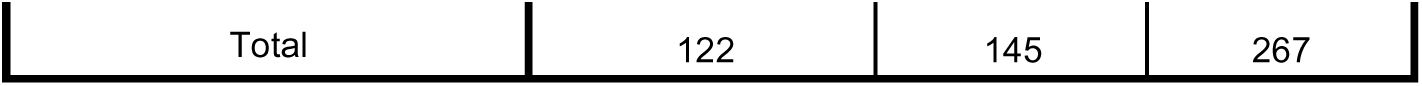
Illustrates the association between the attitude score and academic year.

**Table (4):** Illustrates the association between the knowledge score and attitude score.

**ASCORE2 \* KSCORE2 Crosstabulation**
|  |  | Count |  |  |  |
| --- | --- | --- | --- | --- | --- |
|  |  | KSCORE2 |  |  | Total |
|  |  | POOR | FAIR | GOOD |  |
| ASCORE2 | POOR | 23 | 39 | 37 | 99 |
|  | FAIR | 23 | 46 | 54 | 123 |
|  | GOOD | 6 | 17 | 18 | 41 |
| Total |  | 52 | 102 | 109 | 263 |

n=[(1.96)² x 0.25 x 0.75] / (0.05)² = 288

### Data analysis

Data were compiled and then entered into Microsoft excel. Thus, entered data was transferred to Statistical Package for Social Science (SPSS 24.0 version) for the analysis. P value less than 0.05 was considered significant.

## Results

### socio-demographic characteristics

This study was conducted on 288 respondents, among them 277(96%) subjects have answered the questionnaire, Of which 67(24.2%) were males and 208(75.1%) were females ranging from 17 to 24 years old,most of the subjects were 22 years old (20.9%) . Most of the respondents147(53.1) were medical students, the least of them 58(20.9%) were dental students and the rest the respondents 72(26%) were pharmacy students.

The second year students were 124(44.8%) and the fifth year students 147(53.1%)

### knowledge towards HIV/AIDS among the participants

The total mean score of the study sample on respondents knowledge, attitude and perception(KAP) was found to be 15.45±2.93. which was categorized fair (reference ranging from 13 to 18).

The total mean score of the study sample on respondents knowledge was found to be 7.91±1.69 . which was categorized good (reference ranging from 7 to 10).

More than 97% percent of the subjects responded correctly that HIV is a transmittable disease. More than 77% percent of the subjects responded correctly that HIV is not a hereditary disease.

More than 62% of the respondents know that HIV /AIDS is not curable at this moment .

More than 65% of the subjects know that there is no vaccine for HIV/AIDS until now. More than 96% of the subjects respond correctly that avoiding sharing needles and syringes prevent HIV infections

More than 76% percent of the subjects responded correctly that Having sex with only one faithful, uninfected person prevent HIV infections

More than 68% percent of the subjects responded correctly that avoiding mosquito bites is not necessary to prevent HIV infections

More than 70% of the subjects responded correctly that staying with infected person at the same house does not increase the risk of HIV infections.

More than 70% percent of the subjects responded correctly that using condoms during sexual intercourse prevent HIV infections

More than 95% of the subjects responded correctly that screening donated blood before transfusion prevent HIV infection

### Attitude towards HIV/AIDS among the participants

The total mean score of the study sample on respondents attitude was found to be 2.08±1.22 . which was categorized fair (reference ranging from 2 to 3).

Only 51% of the subject have had a positive attitude regarding comfortable talking with HIV/AIDS patients

More than 48% have had a negative attitude regarding living with people having HIV/AIDS in the same house

Less than half of the respondents 133(48%) indicated that they would be able to work with HIV/AIDS patient

More than 70% of the respondents indicated that HIV/AIDS patients deserve free treatments.

### Perception towards possible causes of HIV infection among the participants

The total mean score of the study sample on respondents attitude was found to be 5.47±1.2 . which was categorized good (reference ranging from 5 to 7).

More than 97% of the subjects responded correctly that HIV can be transmitted by transfusion of HIV infected blood .

More than 90% of the subjects responded correctly that HIV can be transmitted from HIV infected mother to her fetus.

Less than 50% of the subjects responded correctly that HIV can be transmitted by breast feeding from HIV infected mother .

More than 70% of the subjects responded correctly that HIV less likely to be transmitted by kissing an HIV infected person .

More than 74% of the subjects responded correctly that HIV cannot be transmitted by mosquito bites.

More than 90% of the subjects responded correctly that HIV cannot be transmitted by sharing/eating a meal with an HIV infected person.

More than 80% of the subjects responded correctly that HIV cannot be transmitted by Using public toilets.

### Correlation analysis for outcome variables

the attitude score was significantly associated with the gender (p<0.005)

the total knowledge, attitude and perception was significantly associated with the academic year (p<0.000).

the knowledge score was significantly associated with the academic year (p<0.000).

the perception score was significantly associated with the academic year (p<0.000).

no association was found between the attitude score and the academic year(p=0.62) .

no association was found between the attitude score and knowledge score (p=0.736).

## Discussion

The main finding of the current study was satisfactory outcome of the KAP survey in term of evaluating the medical students knowledge, attitude, and perception regarding HIV/AIDS . we have used multiple components questionnaire with diverse domains to level of understanding of medical students feeling,perception to possible causes of HIV infection and their views about possible measures to prevent HIV transmission.

### Knowledge

Most of the subjects have had a clear knowledge about HIV/AIDS, where 41.4% of the respondents have had a good knowledge and more than 38% of the respondents have had a fair knowledge. However, many misconceptions were still noted relating to HIV/AIDS,with 16.7% of the respondents believing that AIDS is a hereditary disease, 19.6% of the subjects believing that HIV/AIDS can be cured at this moment, 13.5% of the respondents believing that there is a vaccine for HIV/AIDS, which was consistently high compared with other results conducted in previous study,(7).,and only 65.8% of them knowing that there is no vaccine for HIV/AIDS, and 18.5% of the respondents believing that avoiding mosquito bites is necessary to prevent HIV infections.. In overall knowledge domain, the subjects have shown a high level knowledge towards HIV/AIDS. However, These fndings are slightly less than the previous studies, (Knowledge and attitude among students towards HIV/AIDS patients at a dental college, Suraram, India). Where 46.8% of the respondents have had an excellent knowledge . also, In consistent with our results, Omidvar and et al in 2000 showed that among nursing student in Babol, only 3.2% of the students had insufficient knowledge about AIDS and 96.8% had good knowledge. the knowledge score was significantly associated with the academic year (p<0.000), where, The overall knowledge level was higher amongst the seniors medical students than their junior counterparts . Surprisingly, there were nearly 19.8% of the respondents have had a poor knowledge on HIV/ AIDS, which was consistently high compared with other results conducted in previous study(KNOWLEDGE AND ATTITUDE TOWARDS HIV/AIDS AMONGST NURSING STUDENTS IN NEPAL).

The results showed no significant association between faculty and student knowledge (p=0.613).However, in consistent with our study a previous study showed a significant difference between faculty and students knowledge (P=0.012) (Knowledge and Attitude in Hamadan University of Medical Sciences Students toward AIDS: A Cross- Sectional Study from West of Iran).

### Attitude

Our study has shown that only 16.5% of the respondents have had a good attitudes, 45.8% of the subjects have had a fair attitudes, and more than 37% of the respondents have had a negative attitudes concerning AIDS and tolerance to HIV patients. it was found that appreciated number of the respondents have expressed less tolerance in talking, willing to live,and working with HIV/AIDS patients, where 28.2% of the subject have had a negative attitude regarding comfortable talking with HIV/AIDS patients and Only 51% of the subject have had a positive attitude regarding comfortable talking with HIV/AIDS patients, More than 49%of the respondents have had a negative attitude regarding living with people having HIV/AIDS in the same house and only 32.6% of the subjects have had a positive attitude regarding living with people having HIV/AIDS in the same house, and Less than half of the respondents 133(48%) indicated that they would be able to work with HIV/AIDS patients and more than 35% of the respondents believed that they would not be able to work with HIV/AIDS patients. However, More than 74% of the respondents indicated that HIV/AIDS patients deserve free treatments.. The results showed no significant association between the academic year and attitude towards HIV/AIDS (p=0.613).However, previous study showed that The overall attitude of senior class was good from their junior counterpart and these attitude differences were found statistically highly significan(KNOWLEDGE AND ATTITUDE TOWARDS HIV/AIDS AMONGST NURSING STUDENTS IN NEPAL). A report from Japan demonstrated a positive change in attitude among college students, (Wang et al.2013). other(Lal et al.2008) study performed on demonstrating a positive association between positive attitude concerning AIDS and tolerance of HIV patients .

We have found only 16.5% of the respondents have had a good attitudes and more than 41% of the respondents have had a good knowledge about HIV/AIDS. Though better knowledge had not shown their changing attitude, we have anticipated that medical students need necessary counselling to pose some influence upon behavioral and attitudinal changes.

### Perception

Most of the subjects have had a clear perception towards the possible causes of HIV infection, where more than 55% of the respondents have had a good perception regarding HIV causes and more than 37% have had a fair perception regarding HIV causes. However, many misconceptions were still noted relating to HIV causes, where more than 37% of respondents believing that HIV cannot be transmitted by breast feeding from an HIV infected mother and only 40.2% responded correctly,more than 23% of the respondents believing that HIV can be transmitted by kissing,more than 18% of the respondents believing that avoiding mosquito bites is necessary to prevent HIV infection. Regarding condom usage, more than 70% of the subjects responded correctly that using condoms during sexual intercourse prevent HIV infections, more than 96% of the subjects responded correctly that avoiding sharing needles and syringes prevent HIV infections and more than 95% of the subjects responded correctly that screening donated blood before transfusion prevent HIV infections. These results were much higher than in n the study by Kumar and co-workers (Goel et al.2010). medical students perception regarding HIV causes is very important in developing prevention intervention strategies.

## Conclusion & Recommendations

### Conclusion

Medical campus students had shown accepted satisfactory levels of knowledge, attitude and perception towards HIV/AIDS. however, many misconceptions were still noted relating HIV/AIDS. better knowledge had not shown their changing attitude, we have anticipated that medical students need necessary counseling to pose some influence upon behavioral and attitudinal changes.

### Recommendations

Medical campus students need necessary counselling to pose some influence upon behavioral and attitudinal changes.

The training programmes for medical students should aim at confidence and skill building to deal with HIV issues.

**Figure (1):**
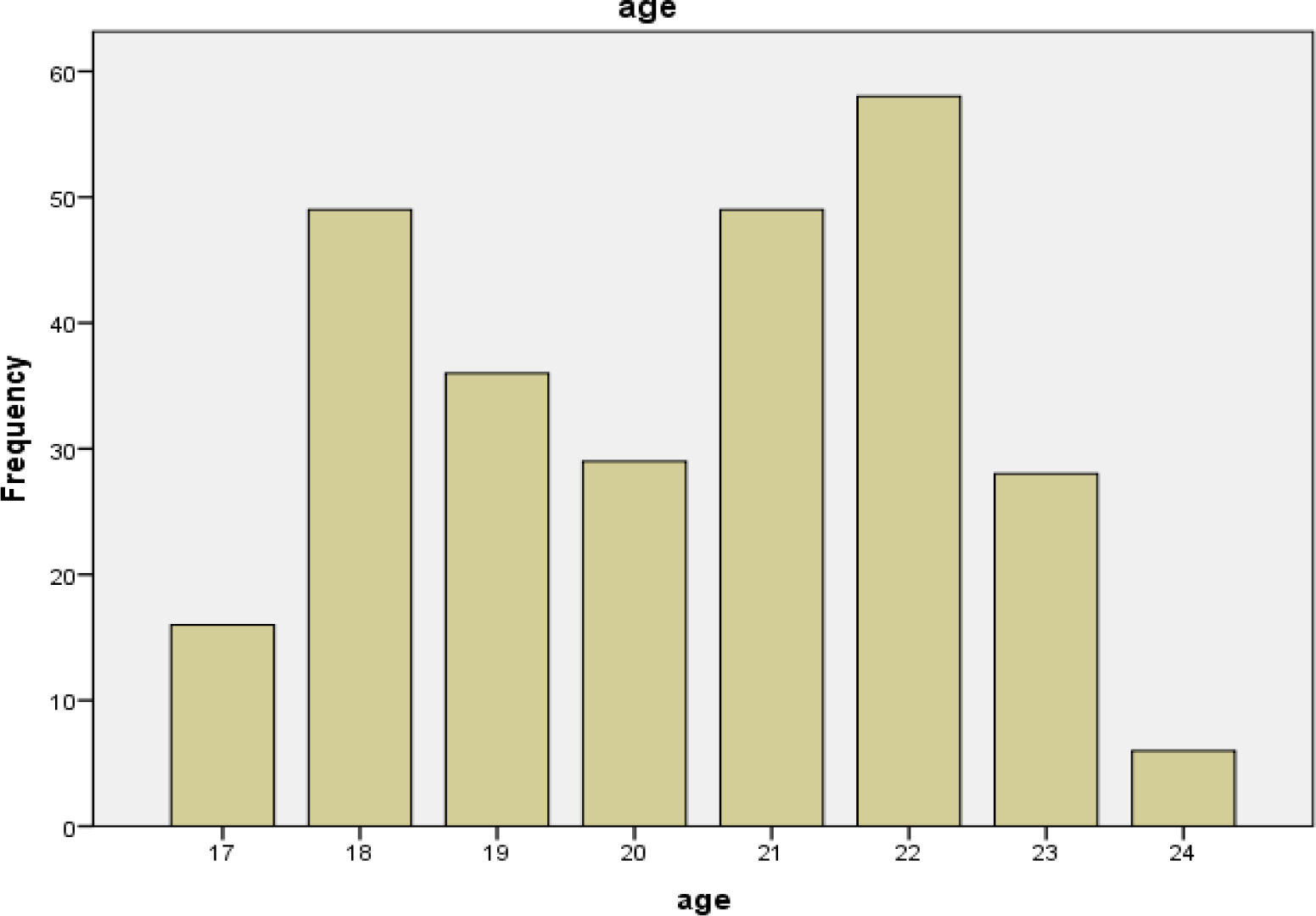
Demographic data (Age)

**Figure (2):**
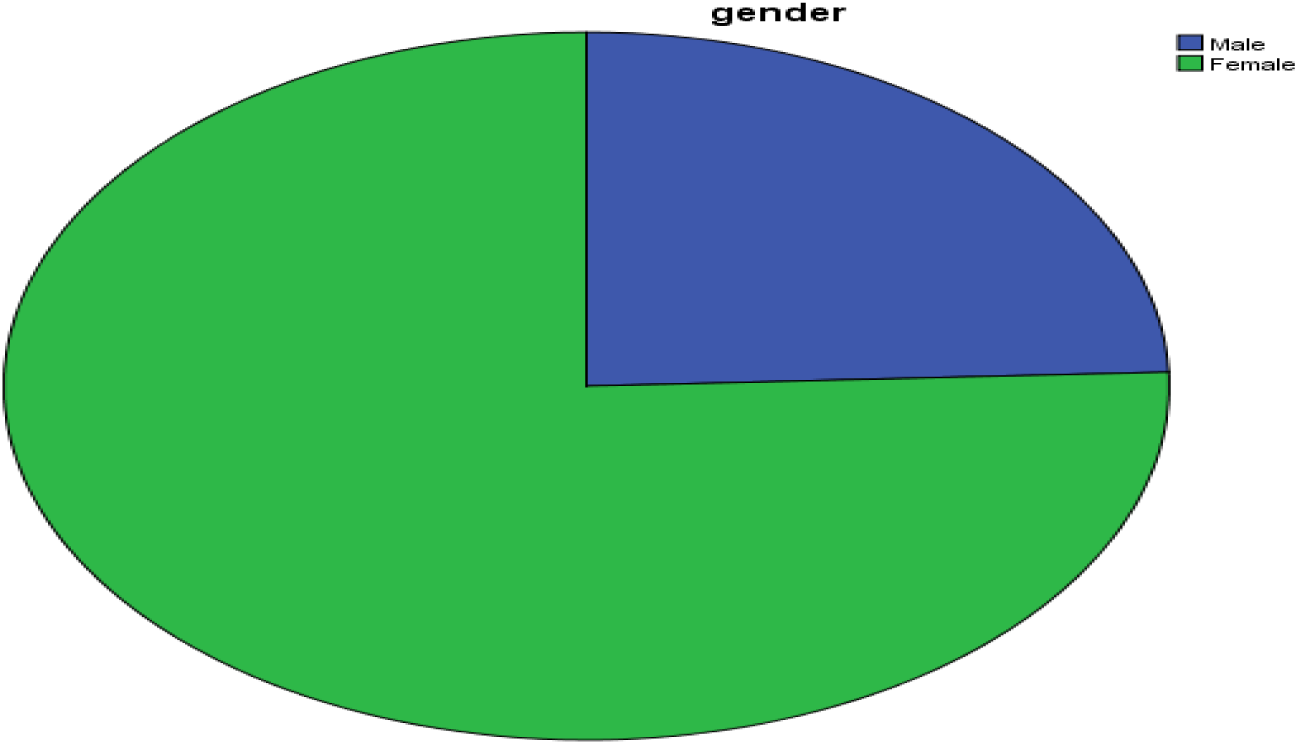
Demographic data (Gender)

**Figure (3):**
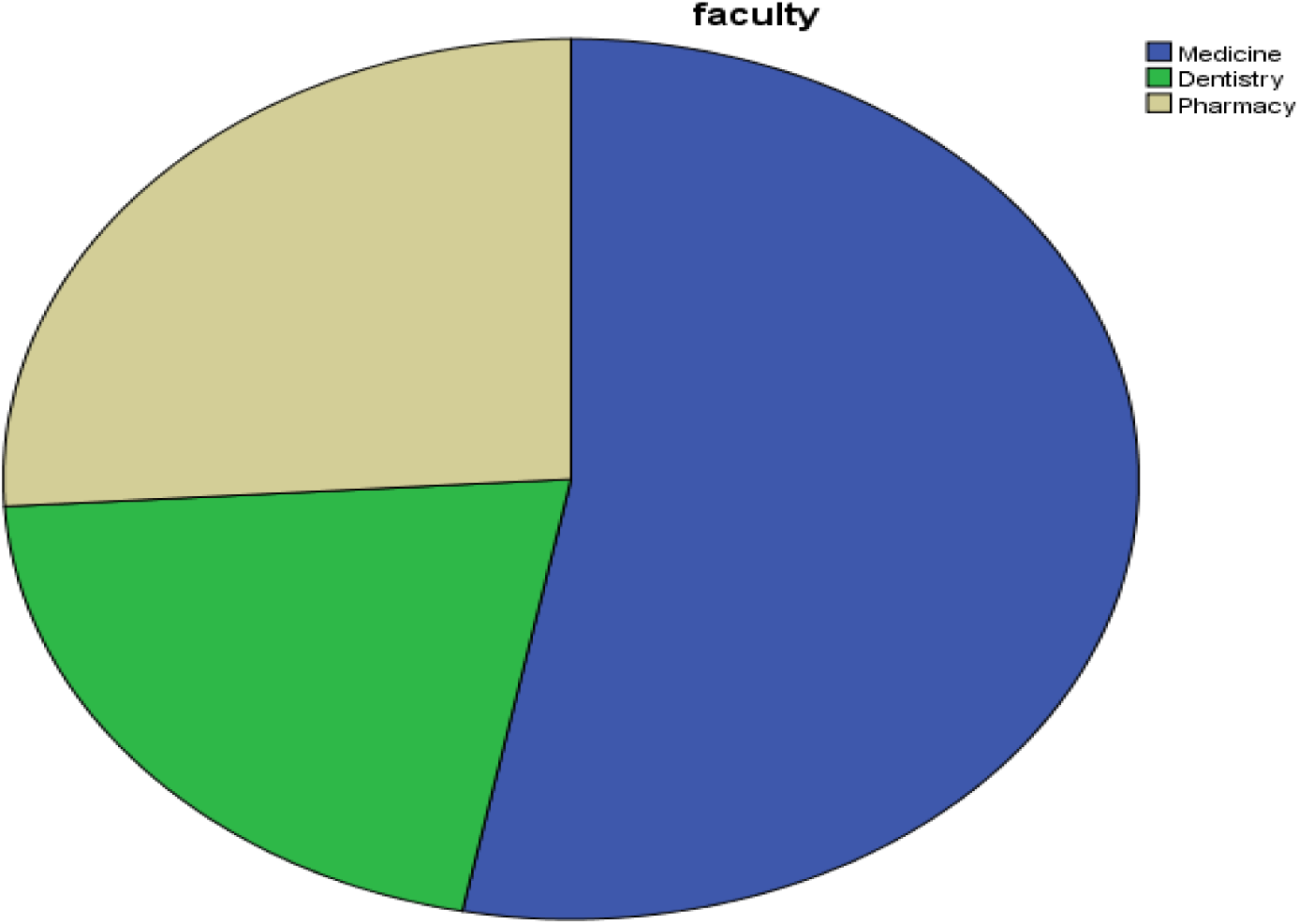
Demographic data (Faculty)

**Figure (4):**
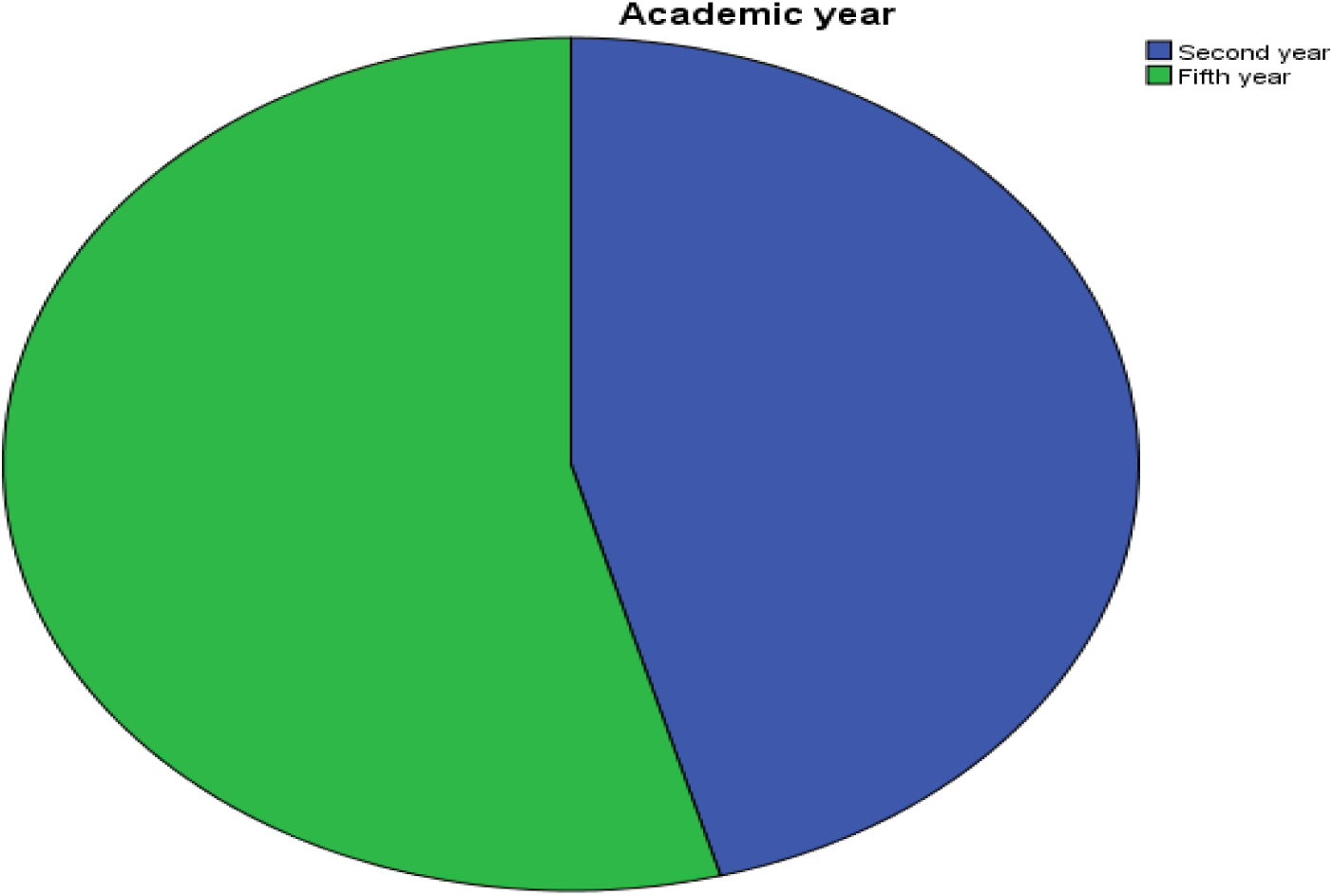
Demographic data (Academic year)

**Figure (5):**
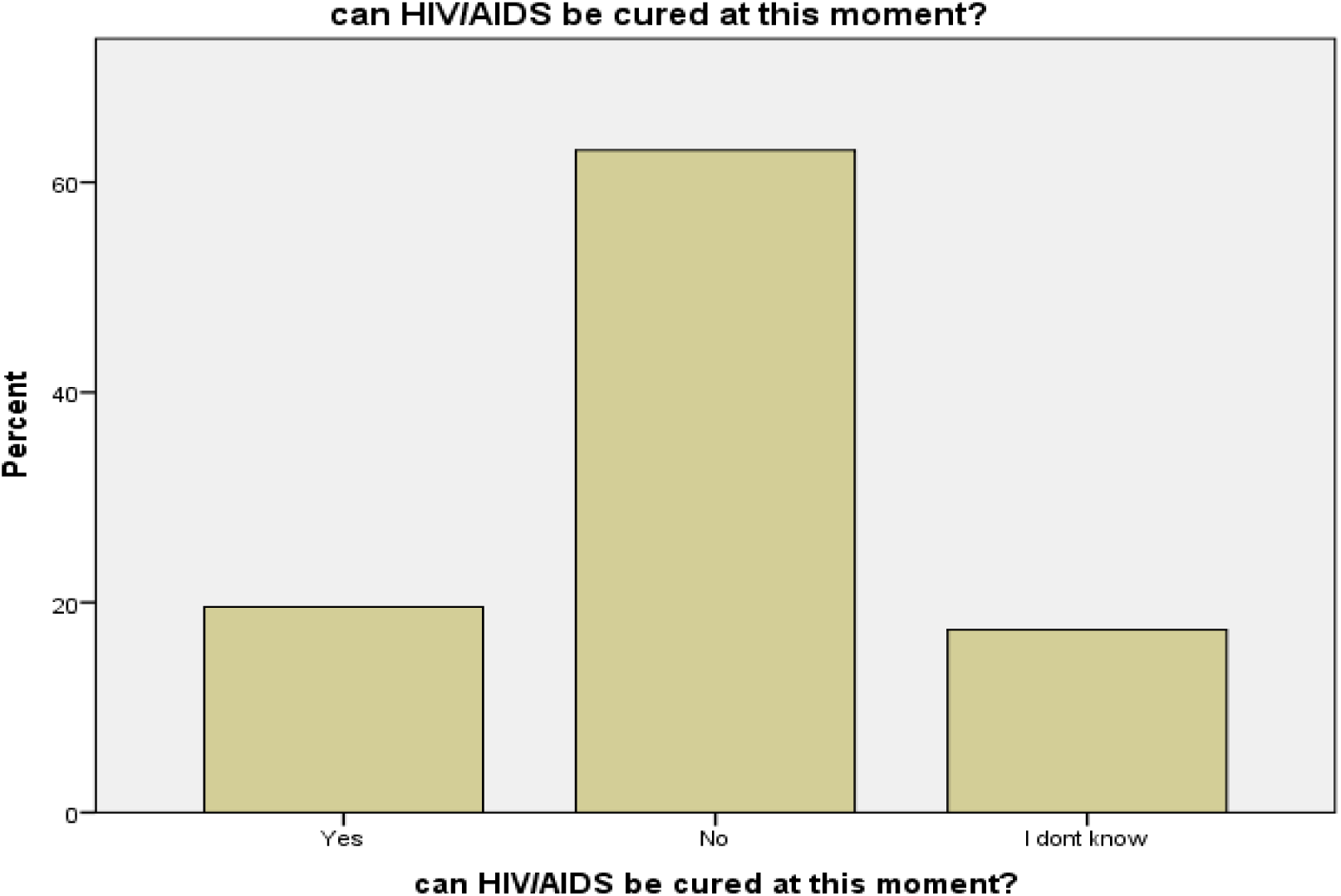
knowledge about AIDS curability.

**Figure (6):**
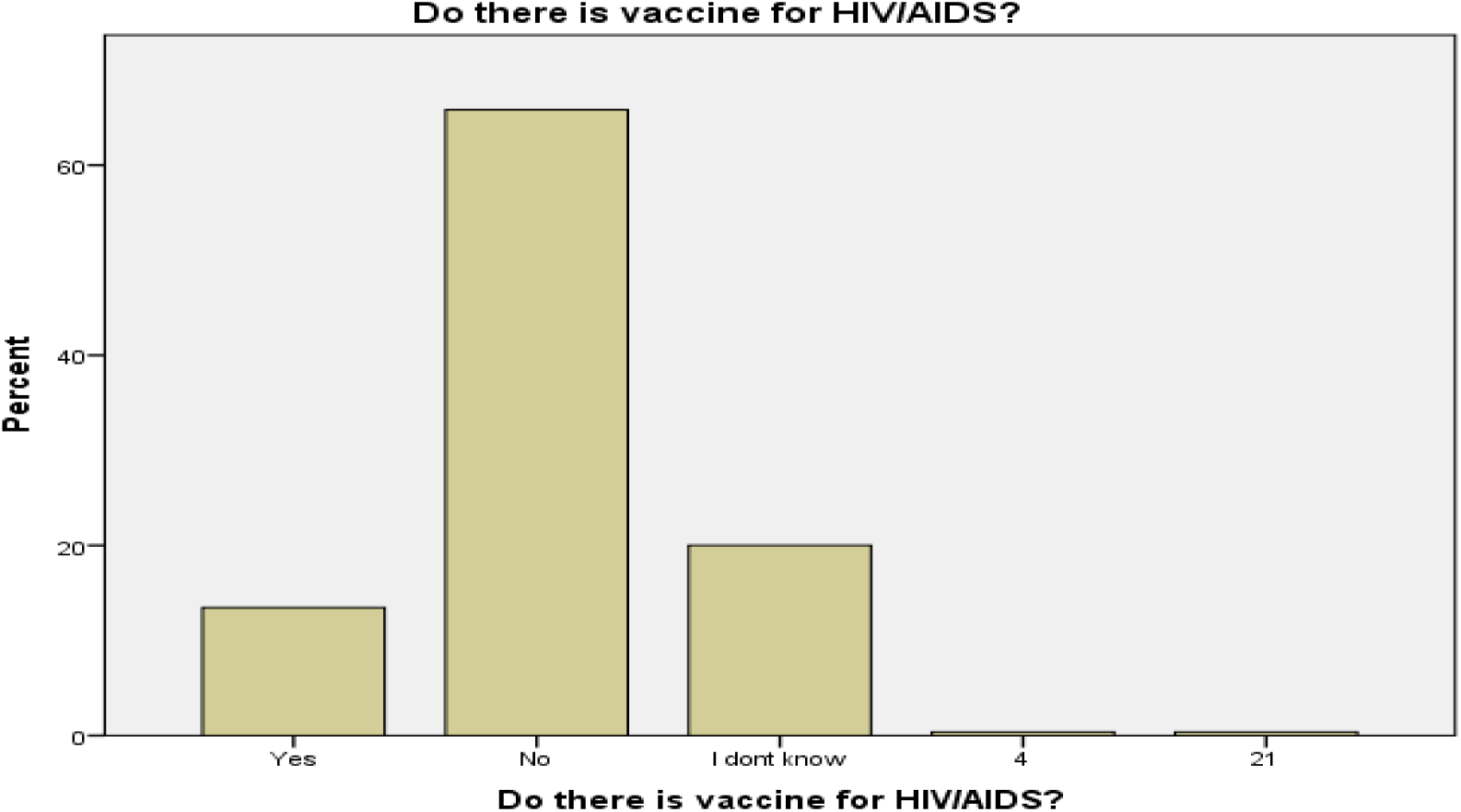
knowledge about HIV vaccine.

**Figure (7):**
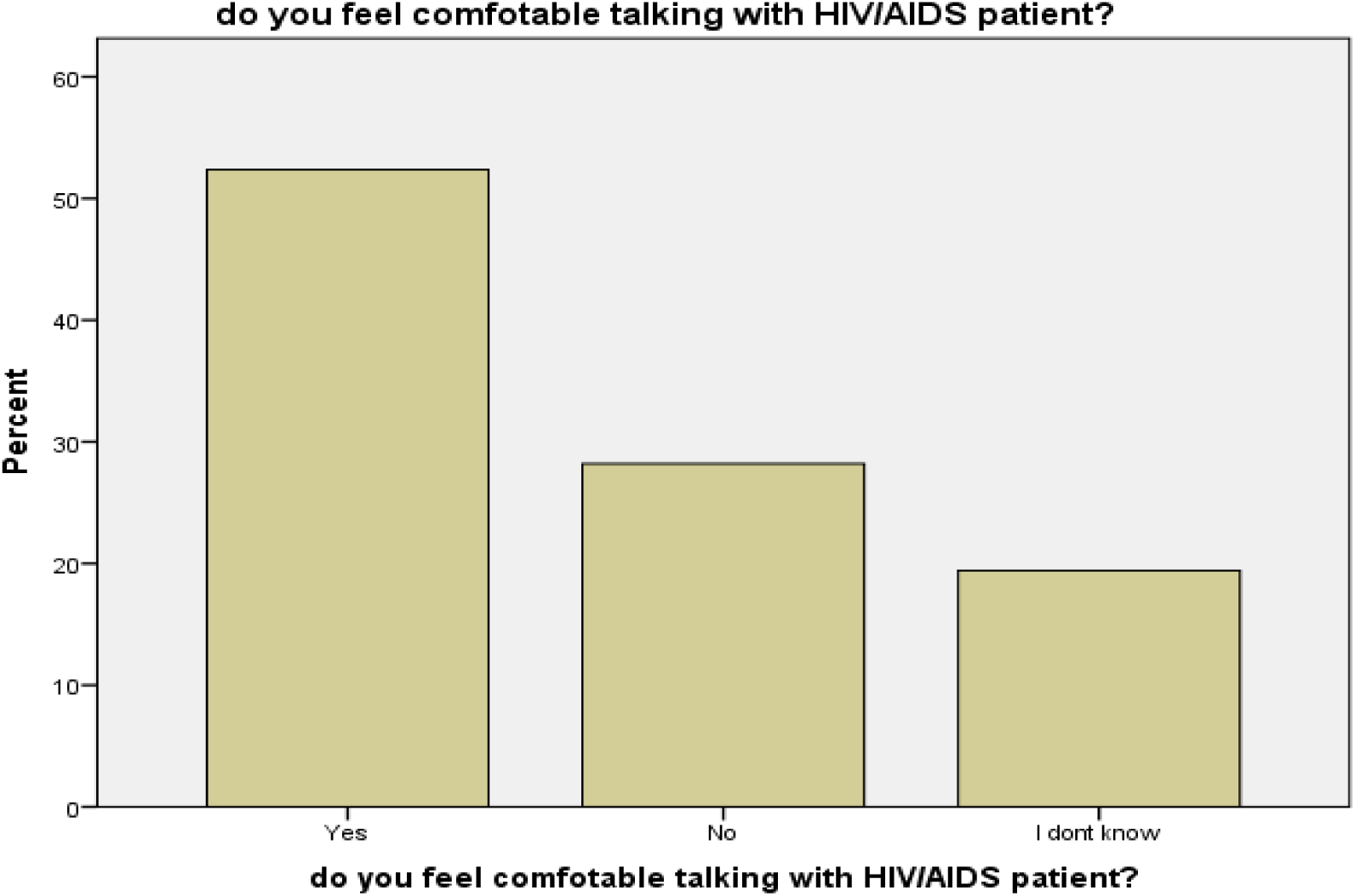
Students attitude towards comfortable talking with AIDS patient

**Figure (8):**
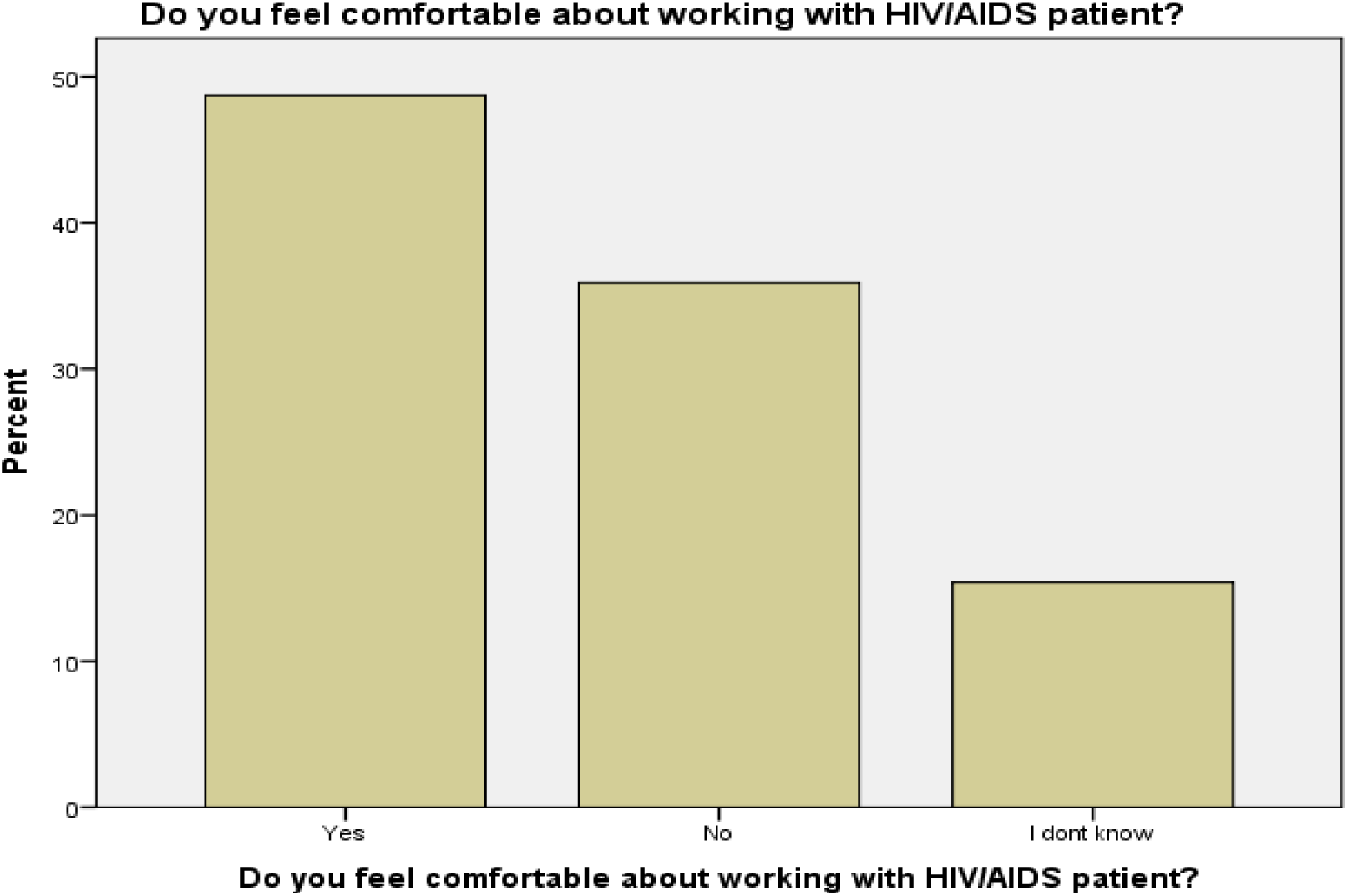
Attitude towards comfortable working with AIDS patients.

**Figure (9):**
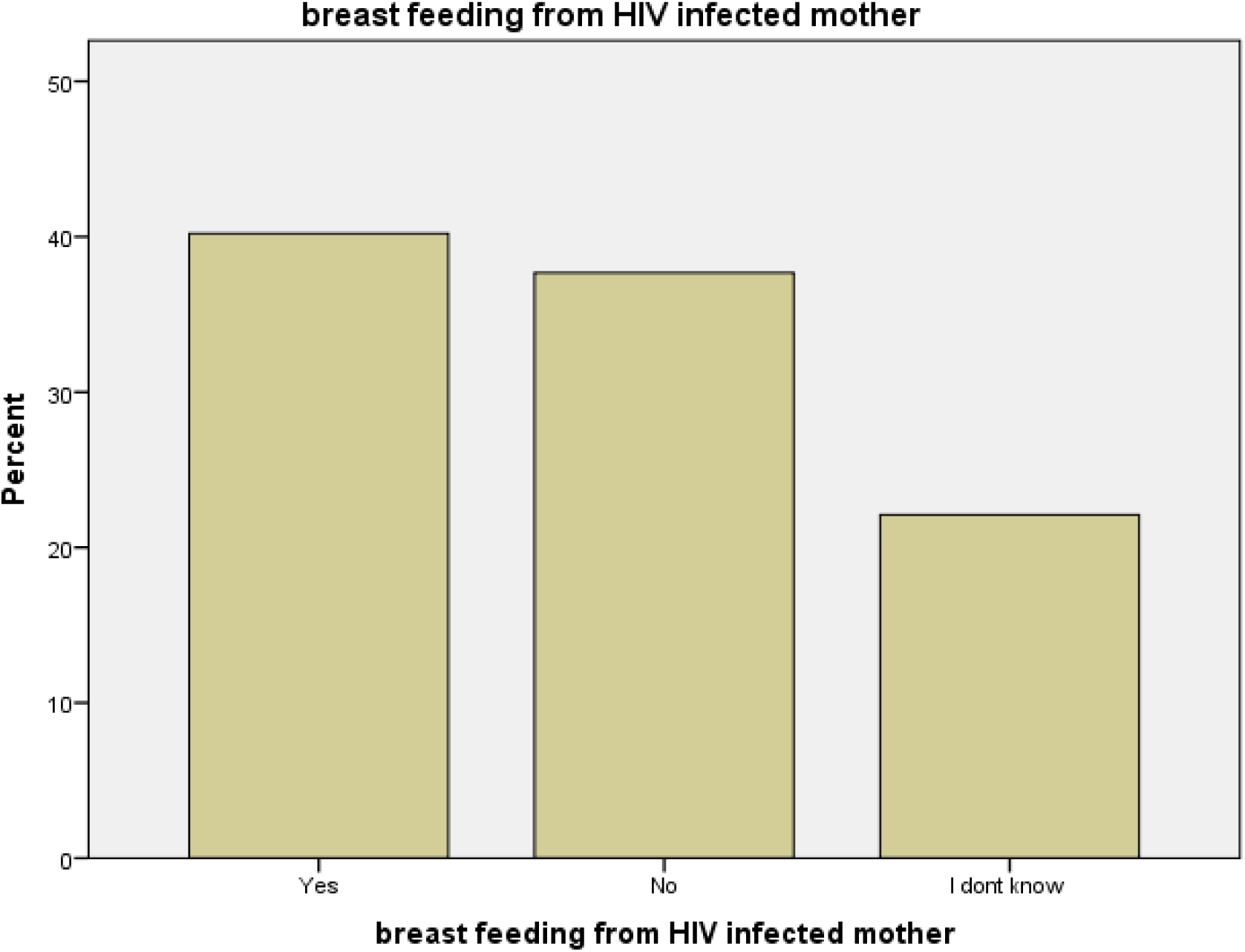
Perception towards breast feeding transmission.

## Data Availability

All data produced in the present work are contained in the manuscript

## List of abbreviations

KAP: knowledge, attitude and perception.
AIDS: Acquired immunodeficiency syndrome.
HIV: Human immunodeficiency virus.
SPSS: Statistical Package for Social Science.

## Declarations

### Ethical approval and consent to participate

Ethical clearance was obtained from the ethical committee of the community medicine department, Faculty of Medicine, University of Khartoum (reference number is not available).

Participants were given detailed information about the study and were asked to provide written consent before completing the questionnaire. Strict measures were implemented to safeguard the privacy and confidentiality of the participants, ensuring that their information remained secure and anonymous.

The study was conducted in line with ethical guidelines and regulations.

### Consent for publication

Not applicable.

### Availability of data and material

The corresponding author can provide the datasets used and analyzed in this study upon reasonable request.

### Author’s contribution

**AA^1^** contributed to the idea, study design, questionnaire design, data collection, data analysis, data interpretation and manuscript drafting.

**RD^2^** contributed to the study design, questionnaire design, data analysis, manuscript drafting.

**RA^3^** contributed to the data collection, data analysis, and manuscript drafting.

**ME^4^** contributed to the data collection and manuscript drafting.

**AA^5^** contributed to manuscript drafting.

**AS^6^** Co-supervisor

**SH^7^** Supervisor

All authors revised the manuscript and approved it for publication.

## Acknowledgment

We express our sincere gratitude to the participants for their exceptional cooperation in this study.

We extend our heartfelt appreciation to the community medicine department at the Faculty of Medicine, University of Khartoum, for their invaluable guidance and support throughout this study.

